# Trends in smoking prevalence and socioeconomic inequalities across regions in England: a population study, 2006 to 2024

**DOI:** 10.1101/2024.10.24.24316046

**Authors:** Sarah E. Jackson, Sharon Cox, Vera Buss, Harry Tattan-Birch, Jamie Brown

**Author notes:** Corresponding author: Dr Sarah Jackson, Department of Behavioural Science and Health, University College London, 1-19 Torrington Place, London WC1E 7HB, UK.

## Abstract

**Background:** In addition to national policies and interventions, certain regions in England (particularly in the North) coordinate regional tobacco control programmes. This study examined trends in tobacco smoking prevalence and socioeconomic inequalities in smoking across regions.

**Methods:** Data were obtained from monthly household surveys of adults (≥16y) in England, conducted between November 2006 and July 2024 (total *n*=368,057). We used logistic regression to estimate time trends in current smoking by region, and tested interactions with occupational social grade to explore differences between more and less advantaged groups.

**Results:** Smoking prevalence declined most in the North (28.8% to 15.8%; -12.9 percentage points [95%CI -14.4; -11.5]), similar to the national average in the Midlands (25.2% to 16.0%; -9.2 [-10.6; -7.9]), and least in the South (22.7% to 17.3%; -5.3 [-6.5; -4.0]), reducing regional disparities such that prevalence was similar across regions in 2024. Socioeconomic inequalities in smoking prevalence between more and less advantaged social grades fell most in Yorkshire and the Humber (from 17.9 percentage points [14.1; 21.8] to 3.7 [0.4; 7.0]) and the West Midlands (from 16.1 [12.8; 19.6] to 3.0 [-0.03; 6.0]). Regions with sustained regional tobacco control activity saw greater declines in smoking prevalence (−18.1 [-21.4; - 14.7]) than regions with none (−12.8 [-13.9; -11.6]).

**Conclusions:** Between 2006 and 2024, smoking rates in the North of England fell faster than the national average, aligning with other regions. Regional tobacco control programmes appeared to contribute to this progress.

## Introduction

In England, there are substantial regional inequalities in health and life expectancy.^1,2^ Across a wide range of health markers, there is evidence of a North-South divide, with the best outcomes in Southern regions of England, the poorest outcomes in Northern regions, and outcomes in the Midlands tending to be closer to the national average.^1,3,4^ Within regions, there are also differences between those who are more and less socioeconomically advantaged.^5,6^

Given tobacco smoking is an important contributor to disease, premature death, and health inequalities,^7–10^ reducing smoking rates has the potential to reduce disparities in health and mortality between and within regions. England has a strong tobacco control climate, with a range of policies (e.g., indoor smoking ban, advertising ban, plain packaging, high taxation) implemented at a national level.^11^ Some regions also undertake dedicated tobacco control activity at a local level, aiming to accelerate reductions in smoking by encouraging and supporting more people to quit.^e.g.,12,13^ Understanding how trends in smoking are changing over time at a regional level can provide insights into best practice. It can also inform the development of targeted interventions to support the government’s tobacco control plan, which aims to reduce inequalities caused and maintained by smoking.^14,15^

Using data collected monthly between 2006 and 2024 as part of a nationally representative survey of adults, this study aimed to estimate time trends in tobacco smoking prevalence in the North, Midlands, and South of England and within each of the nine regions in England, overall and by socioeconomic position (indexed by occupational social grade). A secondary aim was to explore how trends in smoking prevalence have differed between regions with and without dedicated regional tobacco control activity.

## Methods

### Pre-registration

The study protocol and analysis plan were pre-registered on Open Science Framework (https://osf.io/n7kuj/). In addition to our planned analyses of differences in trends across each of the nine regions in England and between regions with sustained vs. no dedicated regional tobacco control activity across the study period, we also analysed differences between the North, Midlands, and South of England.

### Design

Data were drawn from the Smoking Toolkit Study, an ongoing monthly cross-sectional survey representative of adults (≥16 years) in England. Full details of the study’s methodology are available elsewhere.^16,17^ Briefly, the study uses a hybrid of random probability and simple quota sampling to select a new sample of approximately 1,700 adults across England each month. Comparisons with other national surveys and sales data indicate the survey achieves nationally representative estimates of key sociodemographic and smoking variables.^16,18^

The survey began in November 2006 and has been conducted each month since, with the exception of December 2008 and April 2020. Data were collected face-to-face up to the start of the Covid-19 pandemic and via telephone from April 2020 onwards; the two modes show good comparability on key smoking indices.^19^ The lower age limit was raised to 18 years between April 2020 and December 2021.

The present analyses used data collected between November 2006 (the first wave of data collection) and July 2024 (the most recent data available at the time of analysis).

### Measures

Smoking status was assessed by asking participants which of the following best applied to them: (a) I smoke cigarettes (including hand-rolled) every day; (b) I smoke cigarettes (including hand-rolled), but not every day; (c) I do not smoke cigarettes at all, but I do smoke tobacco of some kind (e.g., pipe, cigar or shisha); (d) I have stopped smoking completely in the last year; (e) I stopped smoking completely more than a year ago; or (f) I have never been a smoker (i.e., smoked for a year or more). Those who responded *a* to *c* were considered current smokers.

Time (survey month) was coded from 1 (November 2006) to 213 (July 2024). This coding included months with no data collection; estimates for these months were effectively interpolated at the aggregate level using information before and after the missing time points to model the trends across the period.

Region in England was categorised as North East, North West, Yorkshire and the Humber, East Midlands, West Midlands, East of England, London, South East, and South West. Of these regions, the North East, North West, and Yorkshire and the Humber make up the North of England; the East Midlands, West Midlands, and East of England make up the Midlands; and London, the South East, and the South West make up the South.

We consulted with key stakeholders (Action on Smoking and Health and regional tobacco control coordinators) to determine the provision of regional tobacco control programmes in England across the study period (2006 to 2024; see **Table S1** for a summary of tobacco control activity in each of the nine regions). Between 2005 and 2011, all regions received funding from the Department of Health for tobacco control coordination. During this period, each region had a Regional Tobacco Policy Manager within their Government Office, and from 2008 a Regional Performance and Delivery Manager and Regional Marketing Manager. In the North East, a comprehensive tobacco control programme, ‘Fresh’, was also established in 2005,^12^ benefiting from additional funding from primary care trusts; this is the only region to have additional sustained activity across the entire study period. By contrast, the East Midlands, West Midlands, East of England, and South East have had no additional dedicated regional tobacco control activity. Activity in other regions (the North West, Yorkshire and the Humber, South West, and London) has been inconsistent across the period and varied in intensity between and within regions. We therefore compared trends between regions with sustained (North East) vs. no (East Midlands, West Midlands, East of England, and South East) dedicated tobacco control activity across the study period.

Occupational social grade was categorised based on National Readership Survey classifications^20^ as ABC1 (includes managerial, professional, and upper supervisory occupations) and C2DE (includes manual routine, semi-routine, lower supervisory, state pension, and long-term unemployed). This occupational measure of social grade is a valid index of socioeconomic position that is widely used in research in UK populations and is particularly relevant in the context of tobacco inequalities, use and quitting.^21^

Age was categorised as 16-24, 25-34, 35-44, 45-54, 55-64, and ≥65 years. Gender was self-reported as man, woman, or in another way.

### Statistical analysis

Data were analysed using R v.4.4.1. The Smoking Toolkit Study uses raking to weight the sample to match the population in England.^16^ The following analyses used weighted data (as a sensitivity analysis, we reran one model using unweighted data, which produced a very similar pattern of results; **Figure S1**). We analysed complete cases (with the exception of age and gender, which had a small amount of missing data and were only used to describe the samples surveyed within each region).

We provided descriptive data on sociodemographic characteristics by region. We then used logistic regression to analyse trends in smoking prevalence over the study period, overall (i.e., the national average for England) and by region, among all adults and among those from more and less advantaged social grades. Time was modelled using restricted cubic splines, to allow for flexible and non-linear changes over time, while avoiding categorisation. We analysed differences in trends by three regional variables: (i) the North, Midlands, and South of England; (ii) the nine regions of England; and (iii) regions with sustained vs. no dedicated regional tobacco control activity across the study period. Models of regional trends among all adults included a two-way interaction between time and region, to allow time trends to differ across regions in England. Models of regional trends by social grade included a three-way interaction between time, region, and social grade. For each analysis, we compared models with time analysed using restricted cubic splines with three, four, and five knots (sufficient to accurately model trends across years without overfitting) using the Akaike Information Criterion (AIC). The best fitting model was selected as the model with the lowest AIC or the simplest model within two AIC units (see **Table S2** for details).

We used predicted estimates from the models to plot regional trends in smoking prevalence over the study period alongside the national average trend. We reported absolute percentage point changes in smoking prevalence within each region from the start to the end of the period (November 2006 to July 2024), overall and by social grade. The mean annual change in smoking prevalence can be calculated as the absolute percentage point change across the entire period / 213 monthly waves ^*^ 12. We also calculated the absolute percentage point disparity in smoking prevalence between the less and more advantaged social grades at the start and the end of the period, and the change in the size of this disparity from the start to the end of the period. We reported these alongside 95% confidence intervals (CIs) calculated using bootstrapping with 1,000 replications. We also plotted modelled estimates of (i) the absolute percentage point change in smoking prevalence across the period; and (ii) smoking prevalence in November 2006, September 2015, and July 2024 (the first, middle, and last months in the time series) as heatmaps to display changing patterns of regional variation in smoking prevalence over time.

## Results

A total of 368,912 adults (≥16y) in England were surveyed between November 2006 and July 2024 (mean [SD] = 1,732 [147] per monthly wave). We excluded 855 participants (0.2%) with missing data on smoking status. There were no missing data on region or occupational social grade. Our analysed sample therefore comprised 368,057 participants.

**Table S3** summarises sociodemographic characteristics of the sample, overall and by region.

### Differences between the North, Midlands, and South of England

Figure 1 shows trends in smoking prevalence over the study period in the North, Midlands, and South of England, relative to the national average. **Table 1** presents modelled estimates of smoking prevalence in the first and last months of the study period; **Table 2** provides corresponding estimates by social grade.

**Table 1.** Modelled estimates of smoking prevalence in England in the first and last months of the study period.

|  | Prevalence, % [95%CI]<sup>1</sup> |  | Percentage point change [95%CI]<sup>2</sup> |
| --- | --- | --- | --- |
|  | Nov 2006 | July 2024 |  |
| National average | 25.3 [24.7; 25.9] | 16.5 [16.0; 17.1] | -8.8 [-9.5; -8.0] |
| Region |  |  |  |
| North | 28.8 [27.6; 29.9] | 15.8 [14.9; 16.8] | -12.9 [-14.4; -11.5] |
| North East | 27.4 [25.0; 30.0] | 16.0 [13.7; 18.6] | -11.4 [-14.8; -8.1] |
| North West | 28.3 [26.6; 30.1] | 15.7 [14.3; 17.2] | -12.6 [-14.8; -10.4] |
| Yorkshire and the Humber | 30.0 [28.0; 32.1] | 16.0 [14.4; 17.7] | -14.1 [-16.4; -11.6] |
| Midlands | 25.2 [24.2; 26.3] | 16.0 [15.1; 17.0] | -9.2 [-10.6; -7.9] |
| East Midlands | 26.9 [24.6; 29.2] | 16.4 [14.6; 18.3] | -10.5 [-13.4; -7.8] |
| West Midlands | 26.7 [24.9; 28.6] | 14.7 [13.2; 16.3] | -12.0 [-14.4; -9.6] |
| East of England | 22.9 [21.4; 24.6] | 17.0 [15.4; 18.6] | -6.0 [-8.2; -3.7] |
| South | 22.7 [21.8; 23.6] | 17.3 [16.5; 18.2] | -5.3 [-6.5; -4.0] |
| London | 20.5 [19.1; 21.9] | 17.0 [15.8; 18.4] | -3.4 [-5.4; -1.5] |
| South East | 24.0 [22.5; 25.5] | 16.8 [15.4; 18.2] | -7.2 [-9.2; -5.4] |
| South West | 24.1 [22.1; 26.2] | 18.7 [17.0; 20.6] | -5.3 [-7.7; -2.7] |
| Regional tobacco control activity |  |  |  |
| None<sup>3</sup> | 25.5 [24.8; 26.1] | 16.2 [15.6; 16.7] | -9.3 [-10.0; -8.5] |
| Sustained<sup>4</sup> | 27.6 [25.8; 29.5] | 14.3 [12.8; 16.0] | -13.3 [-15.3; -11.3] |

<sup>1</sup> Data are weighted estimates of prevalence in the first and last months in the study period from logistic regression with survey month modelled non-linearly using restricted cubic splines (with five knots for models comparing trends within regions (3- and 9-level variables) and with three knots for models comparing regions with sustained vs. no regional tobacco control activity; see **Table S2** for model selection).
<sup>2</sup> Percentage point change calculated as prevalence in July 2024 minus prevalence in November 2006 with 95% CIs calculated using bootstrapping (1,000 replications). The mean annual change in smoking prevalence can be calculated as the absolute percentage point change across the entire period / 213 monthly waves \* 12.
<sup>3</sup> East Midlands, West Midlands, East of England, and South East.
<sup>4</sup> North East; note that prevalence estimates differ slightly from the estimates for the North East derived from the model analysing each of the nine regions (although 95%CIs overlap). This is because the best-fitting models for each of these analyses used different numbers of knots to model time.
Corresponding estimates by occupational social grade are provided in **Table 2**.

**Table 2.** Modelled estimates of smoking prevalence in England in the first and last months of the study period, by occupational social grade.

|  | ABC1 (more advantaged) |  |  | C2DE (less advantaged) |  |  |
| --- | --- | --- | --- | --- | --- | --- |
|  | Prevalence, % [95%CI]<sup>1</sup> |  | Percentage point change [95%CI]<sup>2</sup> | Prevalence, % [95%CI]<sup>1</sup> |  | Percentage point change [95%CI]<sup>2</sup> |
|  | Nov 2006 | July 2024 |  | Nov 2006 | July 2024 |  |
| National average | 18.7 [17.9; 19.5] | 12.7 [12.2; 13.3] | -5.9 [-6.8; -5.0] | 33.2 [32.3; 34.1] | 21.4 [20.5; 22.4] | -11.8 [-13.0; -10.4] |
| Region |  |  |  |  |  |  |
| North | 20.8 [19.2; 22.4] | 12.5 [11.5; 13.6] | -8.3 [-10.1; -6.4] | 36.8 [35.2; 38.5] | 19.9 [18.2; 21.6] | -16.9 [-19.2; -14.7] |
| North East | 19.4 [16.3; 23.0] | 10.2 [8.2; 12.8] | -9.2 [-12.9; -5.0] | 36.5 [32.9; 40.2] | 22.7 [18.5; 27.6] | -13.8 [-19.3; -8.3] |
| North West | 21.5 [19.2; 23.9] | 12.2 [10.8; 13.7] | -9.3 [-12.1; -6.8] | 35.7 [33.3; 38.2] | 20.5 [18.1; 23.1] | -15.3 [-18.7; -11.6] |
| Yorkshire and the Humber | 20.5 [17.8; 23.4] | 14.1 [12.3; 16.1] | -6.4 [-9.6; -3.3] | 38.4 [35.6; 41.3] | 17.8 [15.3; 20.6] | -20.6 [-24.2; -16.9] |
| Midlands | 18.1 [16.7; 19.6] | 12.0 [11.1; 13.0] | -6.1 [-7.7; -4.4] | 32.9 [31.4; 34.5] | 20.7 [19.0; 22.4] | -12.3 [-14.6; -9.9] |
| East Midlands | 18.4 [15.6; 21.6] | 11.6 [10.0; 13.5] | -6.8 [-10.1; -3.6] | 35.5 [32.4; 38.9] | 22.0 [18.9; 25.5] | -13.5 [-18.2; -9.2] |
| West Midlands | 18.2 [15.8; 20.9] | 13.0 [11.4; 14.9] | -5.2 [-8.0; -2.2] | 34.3 [31.7; 37.0] | 16.0 [13.6; 18.8] | -18.3 [-21.9; -14.7] |
| East of England | 17.9 [15.9; 20.1] | 11.4 [10.0; 13.0] | -6.5 [-8.8; -4.0] | 29.6 [27.3; 32.1] | 24.5 [21.5; 27.8] | -5.1 [-9.2; -1.4] |
| South | 17.7 [16.6; 18.9] | 13.3 [12.5; 14.1] | -4.5 [-5.9; -3.1] | 30.1 [28.7; 31.6] | 23.2 [21.6; 24.8] | -7.0 [-9.0; -4.9] |
| London | 16.4 [14.7; 18.2] | 13.8 [12.5; 15.2] | -2.6 [-4.7; -0.4] | 26.4 [24.3; 28.7] | 23.2 [20.6; 26.0] | -3.3 [-6.4; +0.4] |
| South East | 18.2 [16.4; 20.1] | 12.2 [11.0; 13.6] | -6.0 [-8.1; -3.8] | 33.7 [31.2; 36.3] | 23.0 [20.5; 25.8] | -10.7 [-14.3; -7.2] |
| South West | 19.1 [16.5; 22.0] | 14.2 [12.4; 16.1] | -5.0 [-8.2; -1.9] | 30.7 [27.7; 33.8] | 23.4 [20.5; 26.6] | -7.3 [-11.2; -2.9] |
| Regional tobacco control activity |  |  |  |  |  |  |
| None<sup>3</sup> | 19.1 [18.3; 19.9] | 12.2 [11.6; 12.8] | -6.9 [-7.7; -6.0] | 34.0 [33.0; 35.0] | 21.1 [20.2; 22.2] | -12.8 [-13.9; -11.6] |
| Sustained<sup>4</sup> | 19.5 [17.2; 22.0] | 9.9 [8.4; 11.6] | -9.6 [-12.1; -7.2] | 37.8 [35.1; 40.6] | 19.7 [17.0; 22.7] | -18.1 [-21.4; -14.7] |

<sup>1</sup> Data are weighted estimates of prevalence in the first and last months in the study period from logistic regression with survey month modelled non-linearly using restricted cubic splines (with five knots for models comparing trends within regions (9- and 3-level variables) and with three knots for models comparing regions with sustained vs. no regional tobacco control activity; see **Table S2** for model selection).
<sup>2</sup> Percentage point change calculated as prevalence in July 2024 minus prevalence in November 2006 with 95% CIs calculated using bootstrapping (1,000 replications). The mean annual change in smoking prevalence can be calculated as the absolute percentage point change across the entire period / 213 monthly waves \* 12.
<sup>3</sup> East Midlands, West Midlands, East of England, and South East.
<sup>4</sup> North East; note that prevalence estimates differ from the estimates for the North East derived from the model analysing each of the nine regions (although 95%CIs overlap). This is because the best-fitting models for each of these analyses used different numbers of knots to model time.

**Figure 1.**
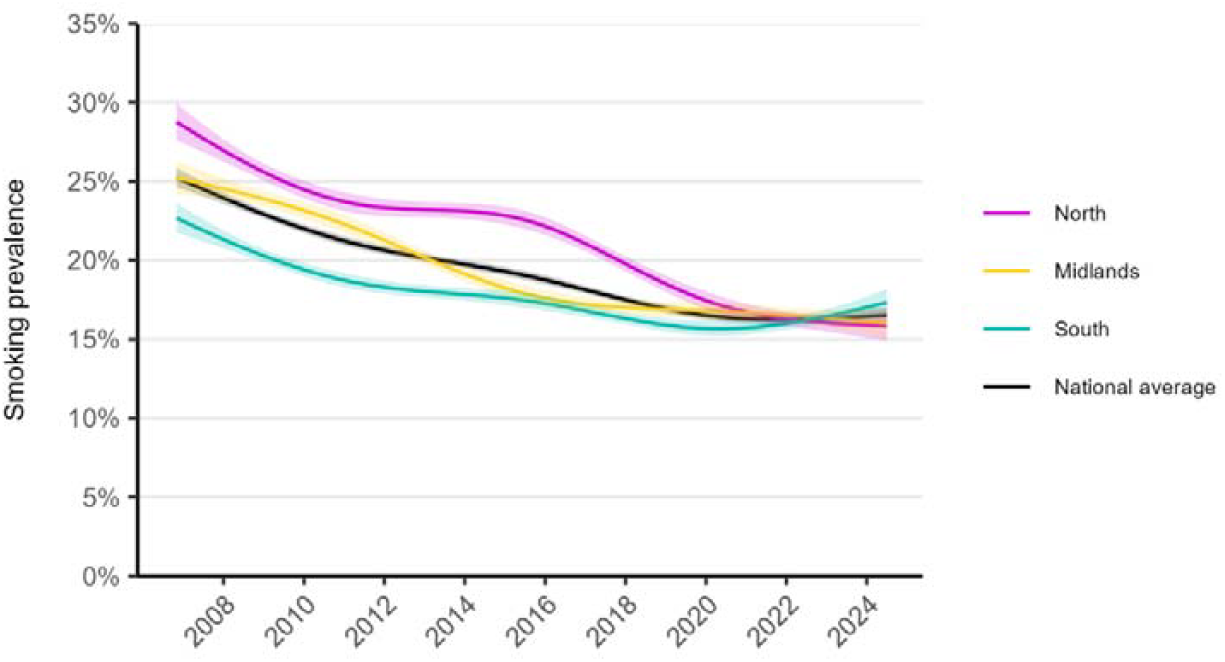
Trends in smoking prevalence in the North, Midlands, and South of England, November 2006 to July 2024. Panels show modelled trends in adult smoking prevalence in the North (North East, North West, and Yorkshire and the Humber), Midlands (East Midlands, West Midlands, and East of England), and South (London, South East, and South West) of England, compared with the national average. Lines represent the modelled weighted prevalence by monthly survey wave (modelled non-linearly using restricted cubic splines with five knots; see **Table S2** for model selection). Shaded bands represent 95% confidence intervals. Unweighted sample sizes: national average *n*=368,057; North *n*=106,174; Midlands *n*=110,004; South *n*=151,879. Corresponding figures by occupational social grade are provided in **Figure S2**.

Overall, smoking prevalence in England declined non-linearly from 25.3% in November 2006 to 16.5% in July 2024; an absolute change of -8.8 percentage points (ppts). Smoking prevalence was consistently higher, but absolute reductions were greater, among those from less advantaged social grades (33.2% to 21.4%; Δ = -11.8 ppts) compared with those who were more advantaged (18.7% to 12.7%; Δ = -5.9 ppts; **Table 2**). As a result, the absolute disparity in smoking prevalence between the more and less advantaged social grades fell from 14.6 ppts in November 2006 to 8.7 ppts in July 2024 (**Table 3**).

**Table 3.** Changes in the absolute disparity in smoking prevalence between more and less advantaged occupational social grades over the study period.

|  | Percentage point disparity<br>[95%CI], C2DE – ABC1<sup>1</sup> |  | Percentage point<br>change [95%CI]<sup>2</sup> |
| --- | --- | --- | --- |
|  | Nov 2006 | July 2024 |  |
| National average | 14.6 [13.4; 15.8] | 8.7 [7.7; 9.9] | -5.9 [-7.3; -4.2] |
| Region |  |  |  |
| North | 16.0 [13.9; 18.4] | 7.4 [5.4; 9.4] | -8.6 [-11.6; -5.8] |
| North East | 17.1 [12.2; 22.0] | 12.4 [7.4; 17.8] | -4.7 [-11.1; 2.5] |
| North West | 14.3 [10.6; 17.7] | 8.3 [5.5; 11.2] | -6.0 [-10.4; -1.7] |
| Yorkshire and the Humber | 17.9 [14.1; 21.8] | 3.7 [0.4; 7.0] | -14.2 [-19.3; -9.3] |
| Midlands | 14.8 [12.6; 17.0] | 8.6 [6.5; 10.7] | -6.2 [-9.2; -3.3] |
| East Midlands | 17.3 [13.0; 21.3] | 10.3 [6.5; 14.2] | -7.0 [-12.5; -1.3] |
| West Midlands | 16.1 [12.8; 19.6] | 3.0 [-0.03; 6.0] | -13.1 [-17.6; -8.5] |
| East of England | 11.8 [8.6; 14.9] | 13.1 [9.7; 16.6] | 1.3 [-3.4; 5.7] |
| South | 12.4 [10.4; 14.2] | 9.9 [8.2; 11.6] | -2.5 [-4.9; -0.02] |
| London | 10.1 [7.3; 13.0] | 9.4 [6.3; 12.3] | -0.7 [-4.6; 3.4] |
| South East | 15.6 [12.4; 18.6] | 10.9 [7.9; 13.9] | -4.7 [-9.2; -0.6] |
| South West | 11.5 [7.4; 15.7] | 9.2 [5.7; 12.7] | -2.3 [-7.4; 3.4] |
| Regional tobacco control activity |  |  |  |
| None<sup>3</sup> | 14.9 [13.5; 16.2] | 9.0 [7.9; 10.1] | -5.9 [-7.3; -4.3] |
| Sustained<sup>4</sup> | 18.3 [14.6; 21.9] | 9.8 [6.6; 13.1] | -8.5 [-12.5; -4.3] |

<sup>1</sup> Data are the difference in weighted estimates of prevalence in the first and last months in the study period between occupational grades C2DE (less advantaged) and ABC1 (more advantaged) with 95% CIs calculated using bootstrapping (1,000 replications).
<sup>2</sup> Percentage point change calculated as percentage point disparity in July 2024 minus percentage point disparity in November 2006 with 95% CIs calculated using bootstrapping (1,000 replications).
<sup>3</sup> East Midlands, West Midlands, East of England, and South East.
<sup>4</sup> North East; note that estimates differ slightly from the estimates for the North East derived from the model analysing each of the nine regions (although 95%CIs overlap). This is because the best-fitting models for each of these analyses used different numbers of knots to model time (see **Table 2**).

At the start of the period, smoking prevalence was highest in the North of England (28.8%), around the national average in the Midlands (25.2%), and lowest in the South (22.7%).

However, over time, the decline in smoking prevalence exceeded the national average in the North (−12.9 ppts), was similar to the national average in the Midlands (−9.2 ppts), and was smallest in the South (−5.3 ppts). This meant that geographic inequalities in smoking prevalence narrowed over time, such that there was little difference in prevalence between the North, Midlands, and South by July 2024 (15.8%, 16.0%, and 17.3%, respectively). This pattern was observed across social grades, although differences in the extent of the decline in smoking prevalence between the North, Midlands, and South were more pronounced in the less advantaged group (−16.9, -12.3, and -7.0 ppts, respectively, compared with -8.3, - 6.1, and -4.5 ppts in the more advantaged group; **Figure S2, Table 2**). This saw the absolute disparity in smoking prevalence between more and less advantaged social grades fall most in the North (from 16.0 to 7.4 ppts), less in the Midlands (14.8 to 8.6 ppts), and least in the South (12.4 to 9.9 ppts).

There was a divergence in trends between 2020 and 2024 (**Figure 1**): a continued decline in smoking prevalence in the North of England (from 17.5% [95%CI 16.9; 18.0] in January 2020 to 15.8% [14.9; 16.8] in July 2024), a small, uncertain decline in the Midlands (16.8% [16.3; 17.4] to 16.0% [15.1; 17.0]), and an increase in the South (15.7% [15.2; 16.1] to 17.3% [16.5; 18.2]). This was driven by differences among the less advantaged social grades; trends between 2020 and 2024 were similar across regions among more advantaged social grades (**Figure S2**).

### Differences across the nine regions of England

Figure 2 shows trends in smoking prevalence over the study period in each of the nine regions of England, relative to the national average; **Figure 3** shows corresponding trends by social grade. **Figure 4** shows modelled estimates of the change in smoking prevalence across the period as heatmaps.

**Figure 2.**
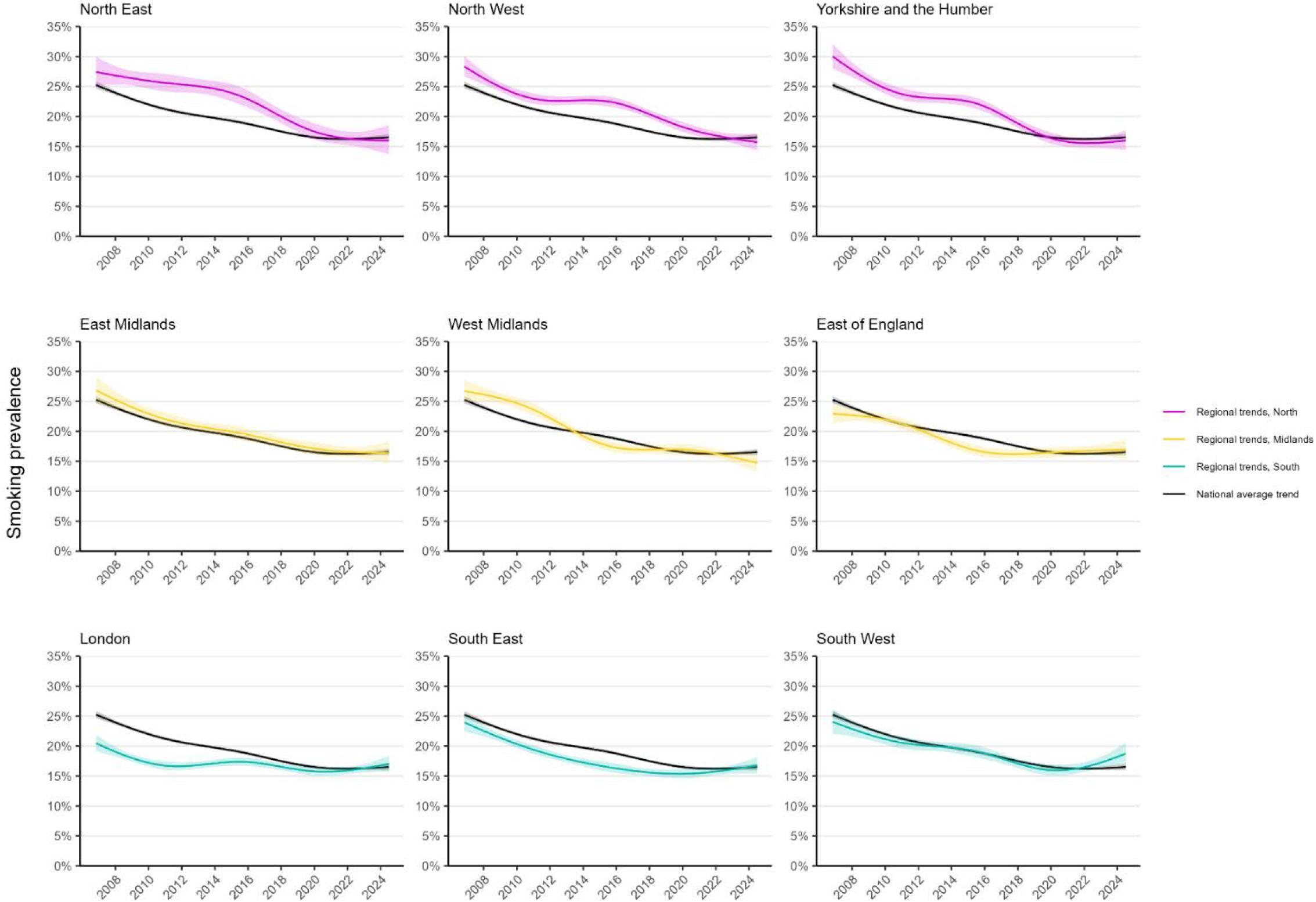
Regional trends in smoking prevalence in England, November 2006 to July 2024. PPanels show modelled trends in adult smoking prevalence in each region in England, compared with the national average. Lines represent the modelled weighted prevalence by monthly survey wave (modelled non-linearly using restricted cubic splines with five knots; see **Table S2** for model selection). Shaded bands represent 95% confidence intervals. Unweighted sample sizes: national average *n*=368,057; North East *n*=18,968; North West *n*=49,584; Yorkshire and the Humber *n*=37,622; East Midlands *n*=30,065; West Midlands *n*=39,030; East of England *n*=40,909; London *n*=61,258; South East *n*=55,367; South West *n*=35,254. Corresponding figures by occupational social grade are provided in **Figure 3**. Trends using unweighted data are shown in **Figure S1**.

**Figure 3.**
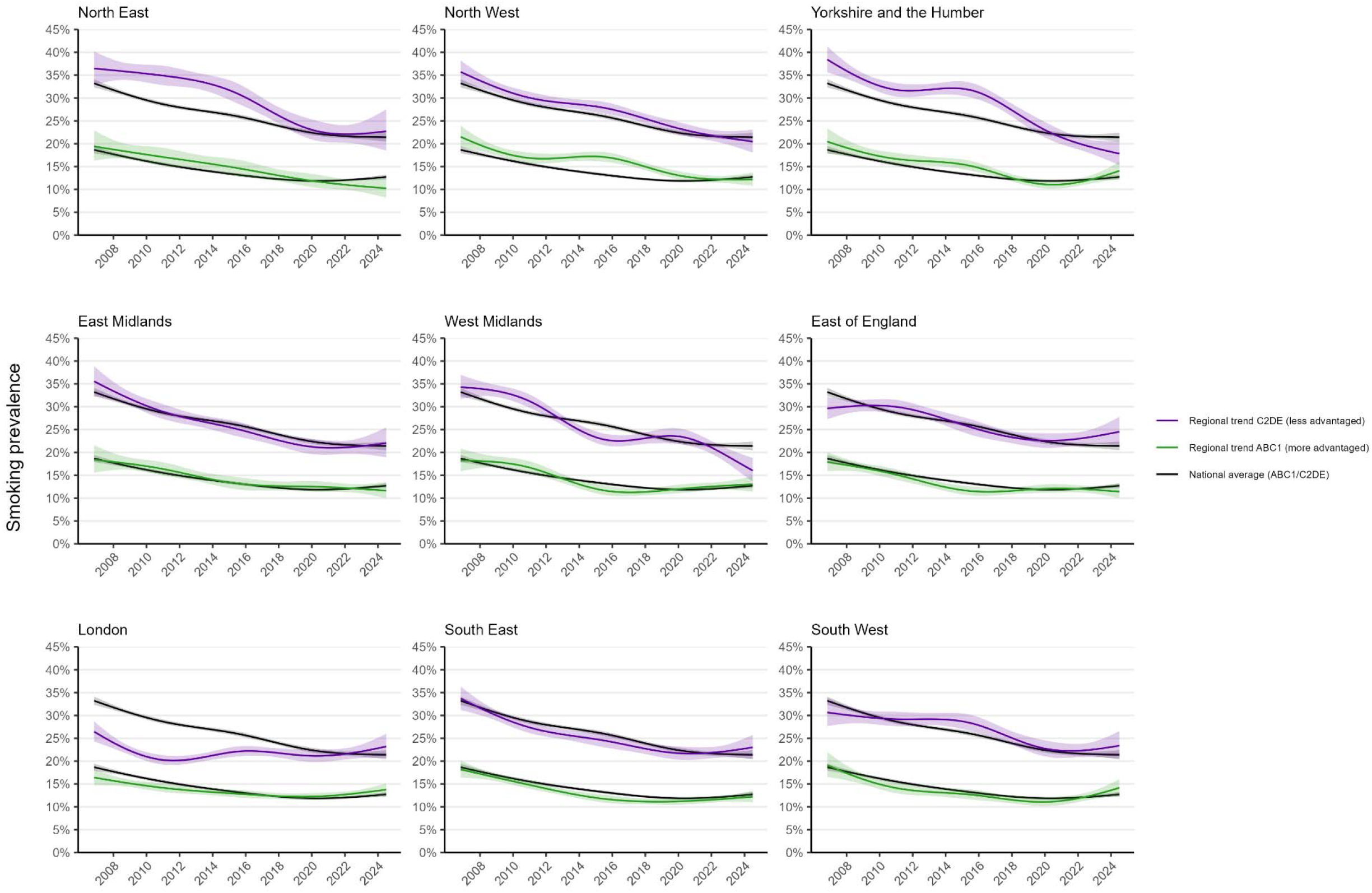
Regional trends in smoking prevalence by occupational social grade in England, November 2006 to July 2024. PPanels show modelled trends in smoking prevalence among adults from more advantaged (ABC1) and less advantaged (C2DE) occupational social grades in each region in England, compared with the national average. Lines represent the modelled weighted prevalence by monthly survey wave (modelled non-linearly using restricted cubic splines with five knots; see **Table S2** for model selection). Shaded bands represent 95% confidence intervals.

**Figure 4.**
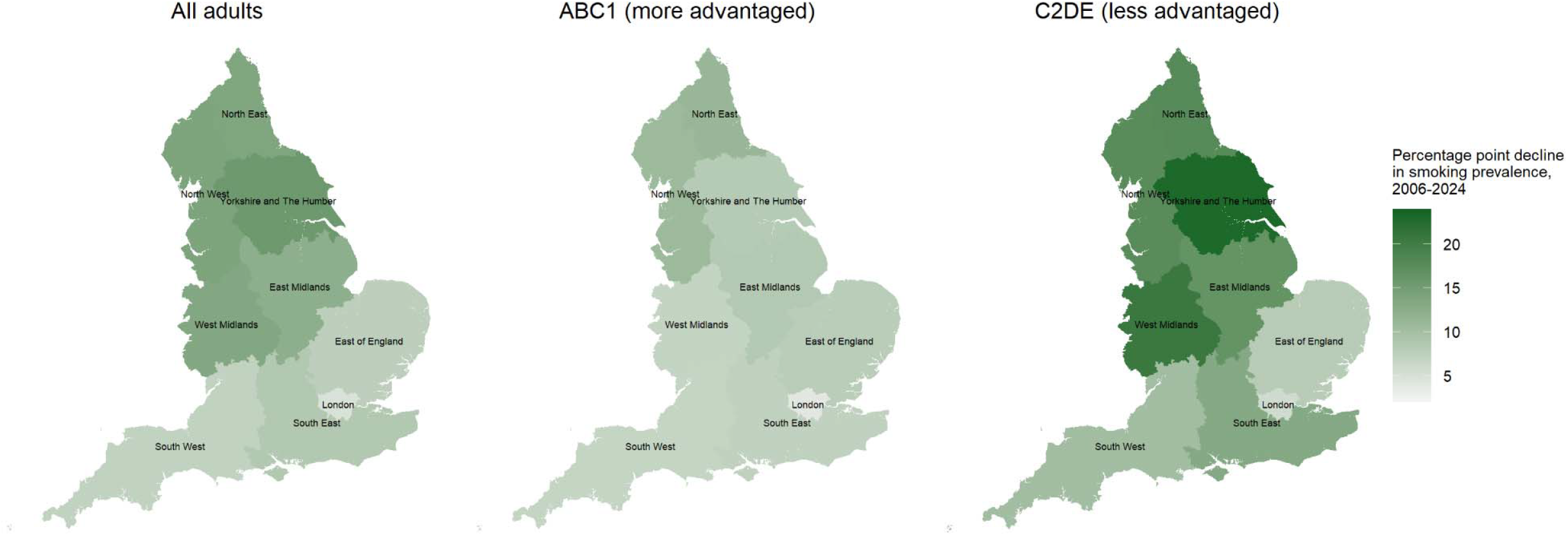
Modelled estimates of changes in smoking prevalence by region in England, November 2006 to July 2024. PPanels show absolute percentage point changes in the modelled weighted estimates of smoking prevalence – among all adults and by occupational social grade – by region in England across the study period. Time (monthly survey wave) was modelled non-linearly using restricted cubic splines with five knots; see **Table S2** for model selection. Heatmaps showing the modelled prevalence of smoking in the first, middle, and last months in the time series are provided in **Figure S3**.

The extent of the decline in smoking was similar across Northern regions but more variable in the Midlands and South. Trends were very similar when modelled using unweighted data (**Figure S1**).

In the North, smoking prevalence was higher than the national average (25.3%) at the start of the period in the North West and Yorkshire and the Humber (28.3% and 30.0%, respectively). It also appeared to be slightly higher in the North East (27.4%) relative to the national average, although there was a slight overlap in the 95%CIs, introducing some uncertainty. The decline in smoking prevalence from the start to the end of the period was greater than the national average decline in each of the Northern regions (−11.4, -12.6, and -14.1 ppts in the North East, North West, and Yorkshire and the Humber compared with the national average of -8.8).

In the Midlands, smoking prevalence was lower than the national average at the start of the period in the East of England (22.9%), compared with the East and West Midlands (26.9% and 26.7%, respectively). However, the change in prevalence from the start to the end of the period was smaller in the East of England (−6.0 ppts) than in the East and West Midlands (−10.5 and -12.0 ppts, respectively), with the decline in smoking having stalled since 2017.

In the South, smoking prevalence in London was substantially below the national average at the start of the period (20.5%) and lower than in the South East or South West (24.0% and 24.1%, respectively). However, the decline in smoking prevalence from the start to the end of the period was smallest in London (−3.4 ppts) and largest in the South East (−7.2 ppts).

Between 2020 and 2024, there was an increase in smoking prevalence in the South West (from 16.0% [15.1; 16.9] in January 2020 to 18.7% [17.0; 20.6] in July 2024) and smaller, uncertain increases in the South East (15.4% [14.7; 16.1] to 16.8% [15.4; 18.2]) and London (15.8% [15.1; 16.5] to 17.0% [15.8; 18.4]).

There were some differences in trends by social grade (**Figure 3, Table 2, Table 3**). Notably, Yorkshire and the Humber and the West Midlands saw the greatest narrowing of inequalities in smoking over time. These regions had large reductions in smoking prevalence among those from less advantaged social grades (−20.6 and -18.3 ppts, respectively), which saw absolute disparities in smoking between less vs. more advantaged groups narrow from 17.9 ppts in November 2006 to 3.7 ppts in July 2024 in Yorkshire and the Humber and from 16.1 ppts to 3.0 ppts in the West Midlands. Alongside the North West, these were the only regions in which smoking prevalence declined in the less advantaged social grades between 2020 and 2024. The pattern of results for London (i.e., lower prevalence at the start of the period and limited change over time) was largely driven by those from less advantaged social grades; the trend for those in more advantaged social grades more closely mirrored the national average.

### Differences between regions with sustained vs. no regional tobacco control activity

Where there was dedicated regional tobacco control activity this was associated with a greater overall decline in smoking (**Figure 5**). Across the period, the total decline in smoking prevalence was -13.3 ppts (−0.75 ppts on average per year) in the region with sustained regional tobacco control activity, compared with -9.3 ppts (−0.52 ppts on average per year) in regions with none (**Table 1**). A similar pattern was observed across those from more and less advantaged social grades (**Figure S4, Table 2**).

**Figure 5.**
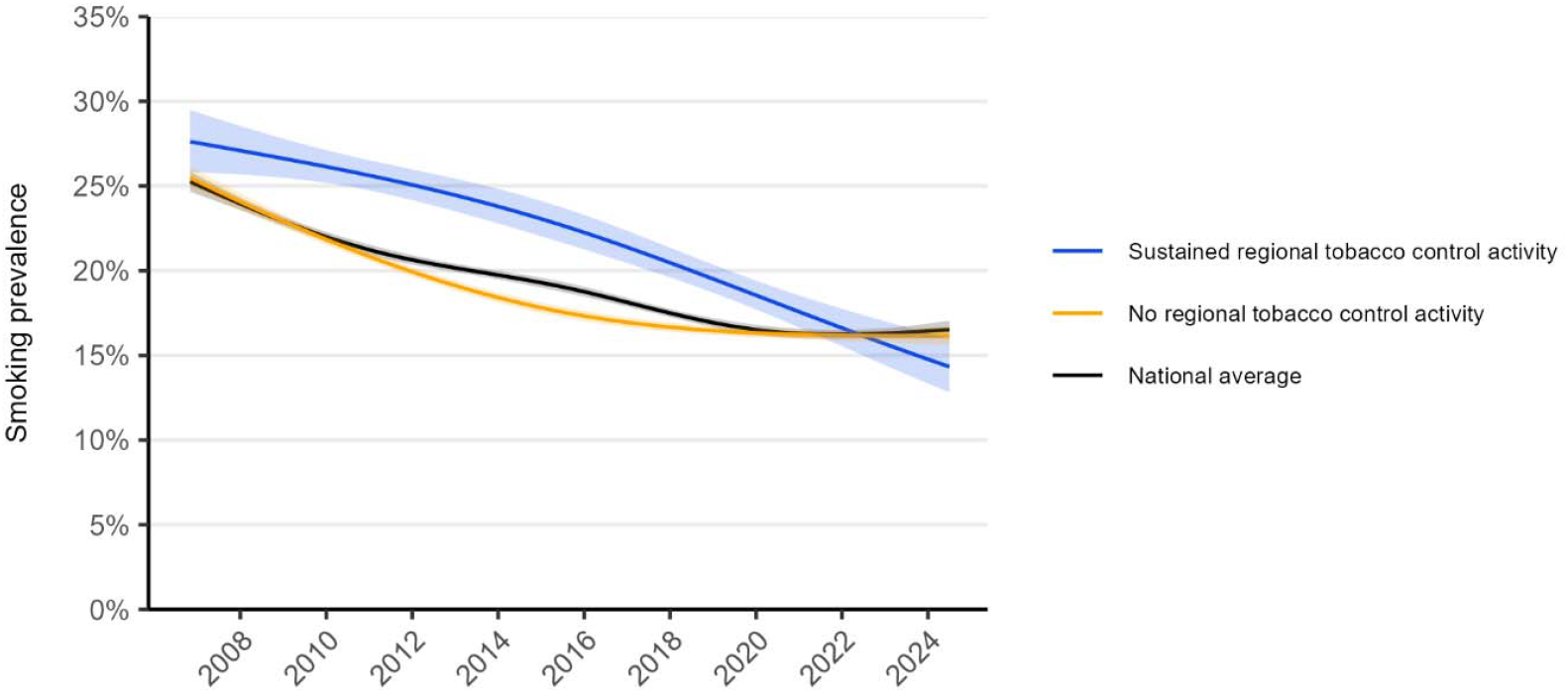
Trends in smoking prevalence in regions in England with sustained vs. no regional tobacco control activity, November 2006 to July 2024. Panels show modelled trends in smoking prevalence among adults in England in regions with sustained regional tobacco control activity and those with no regional tobacco control activity across the study period, compared with the national average. Lines represent the modelled weighted prevalence by monthly survey wave (modelled non-linearly using restricted cubic splines with three knots [regional trends]/five knots [national average trends]; see **Table S2** for model selection). Shaded bands represent 95% confidence intervals. Corresponding figures by occupational social grade are provided in **Figure S4**.

## Discussion

This study provides an overview of regional trends in smoking prevalence in England between 2006 and 2024, with four key findings.

First, regional differences in smoking prevalence narrowed considerably over time. At the start of the period, smoking rates were highest in the North of England and lowest in the South. However, the decline in smoking over time was greatest in the North, similar to the national average in the Midlands, and smallest in the South. As a result, smoking rates have become much more similar across regions in recent years. This broadly mirrors patterns in annual figures reported by region in England’s official estimates of smoking prevalence obtained by the Annual Population Survey up to 2023.^22^ Across this period, socioeconomic inequalities in England have widened,^23,24^ which might be expected to exacerbate inequalities in smoking, but our results show the opposite pattern. This may have important implications for regional health inequalities in decades to come. Smoking is a leading cause of preventable disease and death^10^ and historically, health outcomes have been poorer in the North,^1,2^ where smoking rates were highest.^3^ If they are sustained over time, accelerated declines in smoking may see health outcomes in Northern regions improve and the regional life expectancy gap narrow over time.

Second, differences in trends between regions were particularly pronounced since the start of the Covid-19 pandemic in 2020. A previous analysis of the same dataset indicated the decades-long decline in smoking in England had stalled since the start of the pandemic.^25^ Our present analyses reveal different patterns across the country: smoking prevalence continued to decline in the North but increased in the South (although there was some variation between regions within these broad areas).

Third, regions with sustained dedicated regional tobacco control activity across the period saw greater declines in smoking prevalence than regions with none. Given regional tobacco control programmes have largely been concentrated in the North of England (**Table S1**), this may explain why Northern regions have achieved the greatest reductions in smoking. The North East has had the most consistent investment, coordinating a comprehensive programme since 2005. In other Northern regions, similar programmes have been established at a sub-regional level (e.g., in Greater Manchester within the North West and more recently in Humber and North Yorkshire within Yorkshire and the Humber). By contrast, there has been no dedicated regional tobacco control activity in the Midlands or the South East, and the South West discontinued its regional programme in 2016 due to funding cuts. London is an outlier: despite having some level of coordinated funding since 2014, it had the smallest overall decline in smoking. A previous evaluation of London’s Smoking Cessation Transformation Programme provided evidence of a benefit: the programme launch in 2017 was associated with increased quit attempt rates in the region, over and above changes that occurred in the rest of England, but results were inconclusive regarding an effect on quit success rates.^26^ It is possible the smaller decline in smoking prevalence in London we observed may reflect higher rates of uptake or relapse to smoking in the region (rather than less quitting) or changing population cohorts or demographics in the region (e.g., declines in the average age of people in London or migration from countries with high smoking prevalence).

Fourth, while smoking prevalence was consistently higher among less compared with more advantaged social grades across all regions, some areas have achieved greater reductions in this inequality over time. Socioeconomic disparities in smoking decreased most in Yorkshire and the Humber and the West Midlands, where there were particularly large declines in smoking among less advantaged social grades. In the West Midlands, this brought smoking prevalence among the less advantaged social grades in line with the national average. In London, however, there was little change in smoking prevalence among less advantaged social grades. It is not clear why some regions have achieved better reductions in inequalities in smoking than others – particularly given that one of the regions with the greatest improvement (the West Midlands) is one that has had no dedicated regional tobacco control activity. One possibility is that local authority-commissioned stop smoking services (which exist across all regions of England) are more effective in reaching less advantaged groups in certain parts of the country. It could also potentially be explained by changes in the sociodemographic composition of different social grades within regions over time. Further research is required to better understand the causes of this regional variation and identify best practices that can be applied elsewhere.

Strengths of this study include the large, representative sample and consistent monthly data collection over 17.5 years. National data are also reported by region in England by the Annual Population Survey – crucially the current study is based on monthly estimates to enable trends to be plotted with greater granularity, provides data up to July 2024 and enables an assessment of occupational inequalities within region. However, several limitations should be noted. The observational design means our data can tell us how trends have differed across regions, but not why. While our analyses suggest regional tobacco control programmes may have contributed to the pattern of results we observed, this cannot fully explain why certain regions made greater progress in reducing smoking, and socioeconomic inequalities in smoking, than others. In addition, our analyses cannot identify the mechanisms by which regional tobacco control programmes help to reduce smoking prevalence. Local evaluations may provide important insights into the types of activity that are particularly effective; these findings should be shared nationally to inform allocation of limited resources across regions. Another limitation is the shift from face-to-face to telephone surveys in 2020, though this change affected all regions equally and comparisons suggest the two methods produce similar estimates of smoking prevalence.^19^ Additionally, as a household survey, the data did not capture certain population groups that have high rates of smoking (e.g., people experiencing homelessness or living in institutions); if the size of these groups varies across regions, our results may over- or under-estimate the extent of differences in trends between regions.

In conclusion, between 2006 and 2024, smoking prevalence in the North of England fell faster than the national average, aligning with other regions. Regional tobacco control programmes likely contributed to this progress. The extent to which socioeconomic inequalities in smoking have been reduced has varied across regions.

## Data Availability

Data analysed in the present study are available upon reasonable request to the authors

## Declarations Ethics approval

Ethical approval for the STS was granted originally by the UCL Ethics Committee (ID 0498/001). The data are not collected by UCL and are anonymised when received by UCL.

