## Supplementary material for "Trends in smoking prevalence and socioeconomic inequalities across regions in England: a population study, 2006 to 2024": Table S1

### **Table S1.** Summary of dedicated regional tobacco control activity, 2006 to 2024

| **Region** | **Dedicated regional tobacco control activity** |
| --- | --- |
| North East | Sustained funding (at varying levels over time) for comprehensive *Fresh* programme since 2005 |
| North West | Coordinated region-wide activity between 2007 and 2017 (*Smokefree North West* programme 2007-2010, which became *Tobacco Free Futures* 2010-2017); significant funding invested into sub-regional *Making Smoking History* programme in Greater Manchester since 2017 with spillover across the rest of the region (e.g., media campaigns) |
| Yorkshire and the Humber | Periods of additional funding and coordination across the period, including *Breathe* campaign since 2015; significant funding invested into Humber and North Yorkshire part of the region since 2023 |
| East Midlands | No identified additional regional activity |
| West Midlands | No identified additional regional activity |
| East of England | No identified additional regional activity |
| London | Some level of coordinated funding since around 2014, including the London Smoking Cessation Transformation Programme (which ran *Stop Smoking London*) in 2017-2022 and London Tobacco Alliance and further development of *Stop Smoking London* since 2022 |
| South East | No identified additional regional activity |
| South West | *SmokeFree South West* programme 2008-2016 |

Source: personal communication with Action on Smoking and Health and regional tobacco control coordinators.

### **Table S2.** Model selection: AIC values for models with 3, 4, and 5 knots

|  | **AIC** | | |
| --- | --- | --- | --- |
|  | **3 knots** | **4 knots** | **5 knots** |
| National average trends |  |  |  |
| All adults | 358835.5 | 358834.6 | 358821.6 |
| By social grade | 350412.5 | 350410.9 | 350406.1 |
| Trends within 9 regions of England |  |  |  |
| All adults | 358033.1 | 358035.7 | 357997.2 |
| By social grade | 349709.0 | 349712.8 | 349679.3 |
| Trends within regions with sustained vs. no regional tobacco control activity |  |  |  |
| All adults | 183113.0 | 183113.6 | 183112.8 |
| By social grade | 178435.1 | 178436.5 | 178436.5 |
| Trends within north, midlands,  and south of England |  |  |  |
| All adults | 358177.7 | 358181.5 | 358142.9 |
| By social grade | 349944.9 | 349953.5 | 349916.5 |

AIC, Akaike Information Criterion.

Shaded cells indicate the best fitting model (the model with the lowest AIC or the simplest model within 2 AIC units).

### **Table S3.** Sample characteristics by region (data aggregated across the study period)

|  | **Overall** | **North East** | **North West** | **Yorkshire and the Humber** | **East Midlands** | **West Midlands** | **East of England** | **London** | **South East** | **South West** |
| --- | --- | --- | --- | --- | --- | --- | --- | --- | --- | --- |
| **N** | 368,057 | 18,968 | 49,584 | 37,622 | 30,065 | 39,030 | 40,909 | 61,258 | 55,367 | 35,254 |
| **Age (years)** |  |  |  |  |  |  |  |  |  |  |
| 18-24 | 14.3 | 13.9 | 16.5 | 14.8 | 14.1 | 15.9 | 12.4 | 16.7 | 12.0 | 11.7 |
| 25-34 | 16.5 | 15.8 | 15.7 | 16.6 | 16.1 | 16.6 | 15.2 | 23.4 | 14.1 | 13.3 |
| 35-44 | 17.0 | 16.5 | 15.7 | 16.6 | 16.5 | 16.4 | 17.0 | 20.5 | 17.0 | 15.2 |
| 45-54 | 16.7 | 17.5 | 16.2 | 16.7 | 16.8 | 15.9 | 16.9 | 16.0 | 17.9 | 17.0 |
| 55-64 | 14.4 | 16.1 | 14.6 | 15.1 | 15.1 | 14.3 | 15.4 | 10.7 | 15.1 | 15.8 |
| 65+ | 21.0 | 20.3 | 21.3 | 20.3 | 21.4 | 20.9 | 23.0 | 12.7 | 23.9 | 27.0 |
| Missing, *n* | 101 | 1 | 7 | 11 | 10 | 11 | 15 | 23 | 13 | 10 |
| **Gender** |  |  |  |  |  |  |  |  |  |  |
| Men | 48.8 | 47.9 | 48.7 | 48.5 | 49.0 | 48.7 | 48.5 | 50.5 | 48.3 | 48.0 |
| Women | 51.1 | 52.0 | 51.2 | 51.3 | 50.8 | 51.1 | 51.4 | 49.3 | 51.5 | 51.8 |
| Other | 0.2 | 0.2 | 0.2 | 0.2 | 0.2 | 0.2 | 0.1 | 0.2 | 0.2 | 0.1 |
| Missing, *n* | 391 | 9 | 45 | 44 | 34 | 37 | 43 | 89 | 53 | 37 |
| **Social grade** |  |  |  |  |  |  |  |  |  |  |
| AB (most advantaged) | 26.9 | 22.9 | 23.6 | 26.6 | 22.8 | 23.9 | 29.7 | 27.0 | 31.1 | 30.2 |
| C1 | 28.7 | 27.4 | 28.2 | 27.5 | 27.2 | 27.6 | 29.4 | 30.8 | 30.6 | 27.0 |
| C2 | 20.9 | 22.3 | 21.8 | 20.7 | 24.3 | 21.1 | 20.3 | 18.7 | 20.5 | 21.0 |
| D | 14.9 | 16.0 | 15.1 | 16.1 | 17.5 | 17.8 | 13.4 | 15.0 | 11.9 | 14.4 |
| E (least advantaged) | 8.5 | 11.4 | 11.3 | 9.1 | 8.2 | 9.5 | 7.1 | 8.6 | 6.0 | 7.5 |
| Missing, *n* | 0 | 0 | 0 | 0 | 0 | 0 | 0 | 0 | 0 | 0 |

Data shown are weighted percentages, unless otherwise specified. Sample sizes and numbers of missing cases are presented as unweighted *n*s.

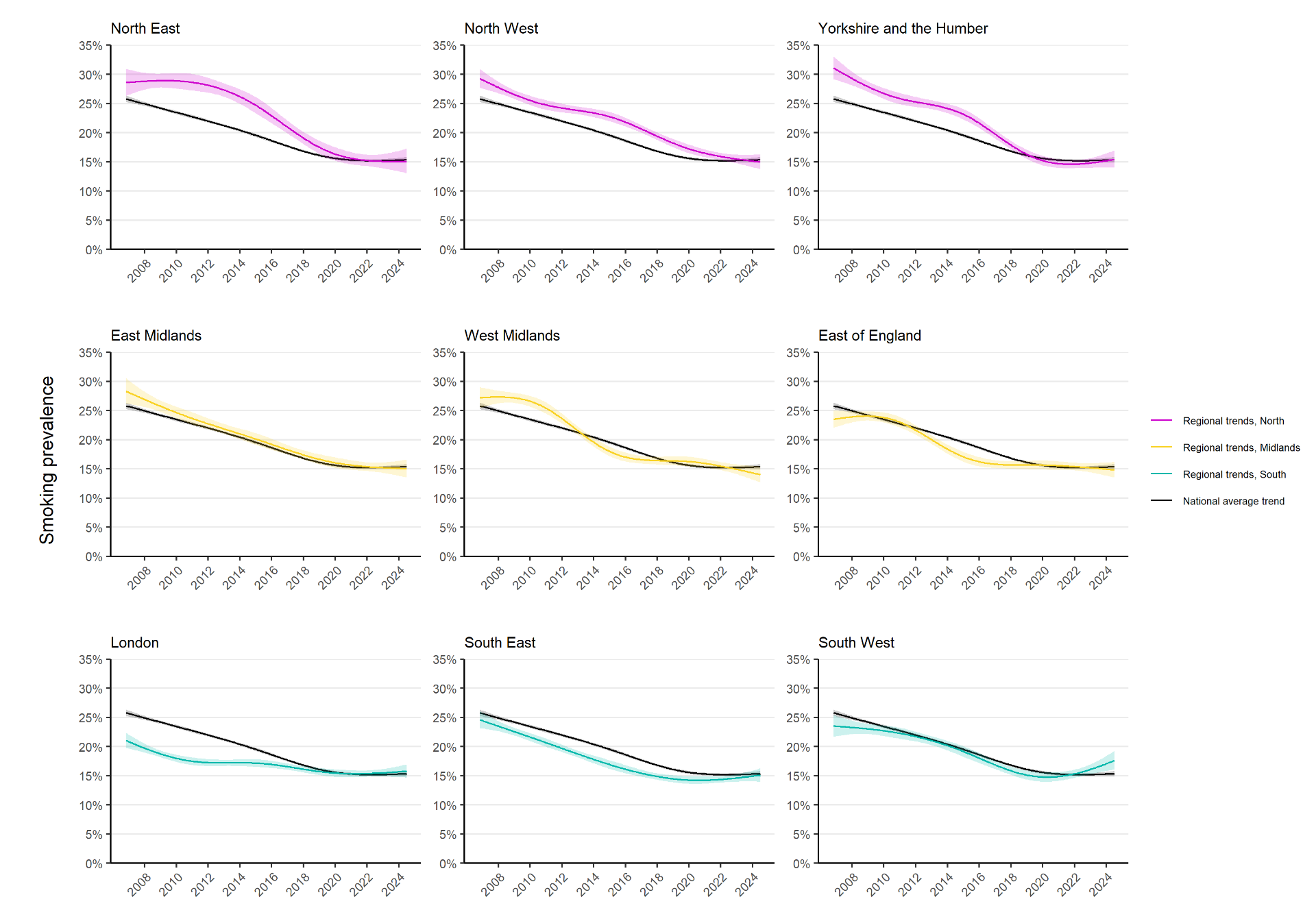

### **Figure S1. Regional trends in smoking prevalence in England, November 2006 to July 2024 (unweighted data).** Panels show modelled trends in adult smoking prevalence in each region in England, compared with the national average. Lines represent the modelled unweighted prevalence by monthly survey wave (modelled non-linearly using restricted cubic splines with five knots). Shaded bands represent 95% confidence intervals. Sample sizes: national average n=368,057; North East n=18,968; North West n=49,584; Yorkshire and the Humber n=37,622; East Midlands n=30,065; West Midlands n=39,030; East of England n=40,909; London n=61,258; South East n=55,367; South West n=35,254.

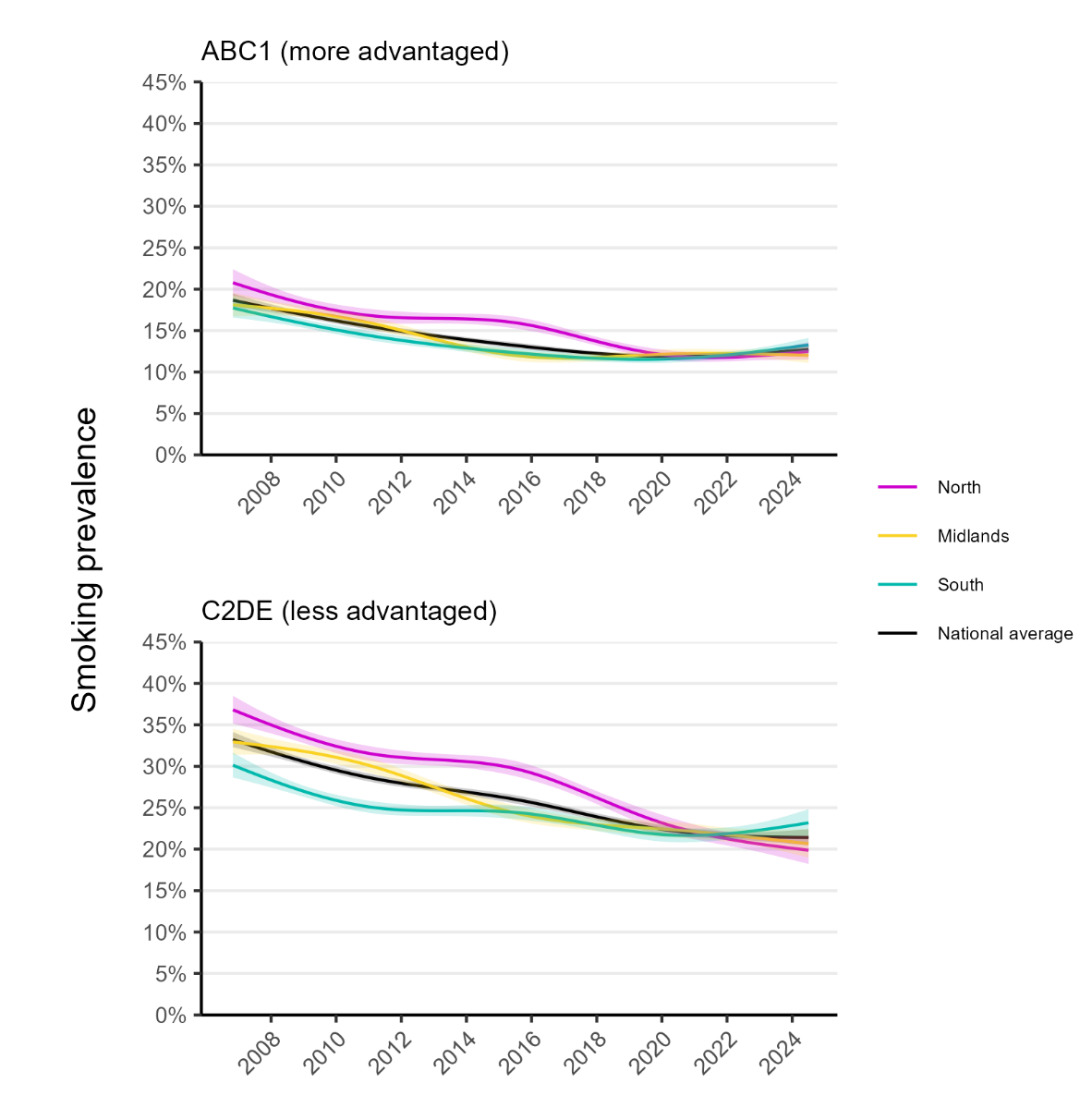

### **Figure S2.** **Trends in smoking prevalence in the north, midlands, and south of England, November 2006 to July 2024.** Panels show modelled trends in smoking prevalence by occupational social grade in the north (North East, North West, and Yorkshire and the Humber), midlands (East Midlands, West Midlands, and East of England), and south (London, South East, and South West) of England, compared with the national average. Lines represent the modelled weighted prevalence by monthly survey wave (modelled non-linearly using restricted cubic splines with five knots; see **Table S2** for model selection). Shaded bands represent 95% confidence intervals.

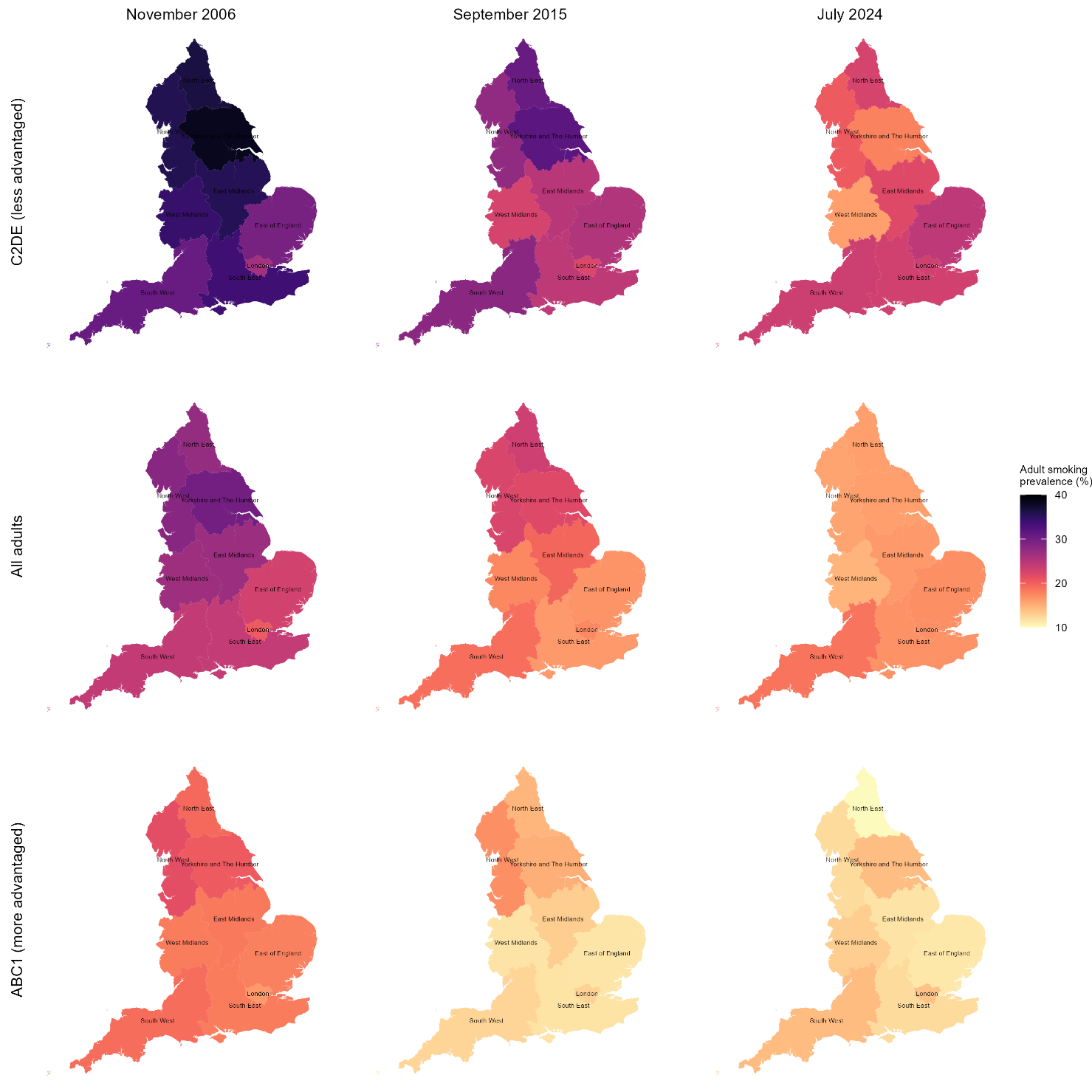

### **Figure S3. Modelled estimates of smoking prevalence by region in England at the start, middle, and end of the study period.** Panels show modelled weighted estimates of smoking prevalence – among all adults and by occupational social grade – by region in England in November 2006, September 2015, and July 2024 (the first, middle, and last months of the study period). Time (monthly survey wave) was modelled non-linearly using restricted cubic splines with five knots; see **Table S2** for model selection.

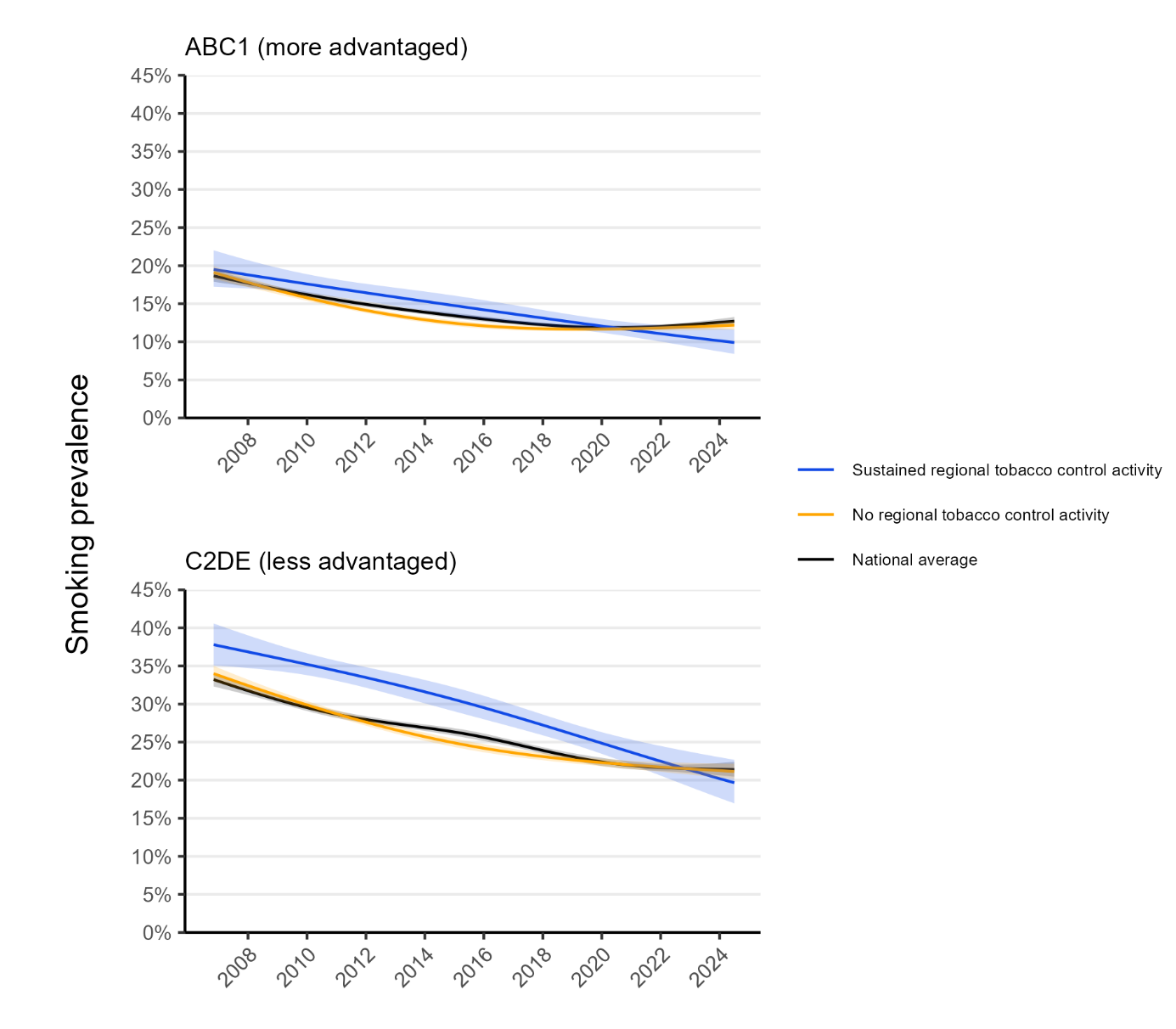

### **Figure S4.** **Trends in smoking prevalence in regions in England with sustained vs. no regional tobacco control activity, November 2006 to July 2024.** Panels show modelled trends in smoking prevalence by occupational social grade in regions with sustained regional tobacco control activity and those with no regional tobacco control activity across the study period, compared with the national average. Lines represent the modelled weighted prevalence by monthly survey wave (modelled non-linearly using restricted cubic splines with three knots [regional trends]/five knots [national average trends]; see **Table S2** for model selection). Shaded bands represent 95% confidence intervals.
